# Deep learning-based automatic classification of ischemic stroke subtype using diffusion-weighted images

**DOI:** 10.1101/2024.02.02.24302247

**Authors:** Wi-Sun Ryu, Dawid Schellingerhout, Hoyoun Lee, Keon-Joo Lee, Chi Kyung Kim, Beom Joon Kim, Jong-Won Chung, Jae-Sung Lim, Joon-Tae Kim, Dae-Hyun Kim, Jae-Kwan Cha, Leonard Sunwoo, Dongmin Kim, Sang-Il Suh, Oh Young Bang, Hee-Joon Bae, Dong-Eog Kim

**Affiliations:** Department of Neurology, Dongguk University Ilsan Hospital, Goyang, Republic of Korea; Artificial Intelligence Research Center, JLK Inc., Seoul, Republic of Korea; Department of Neuroradiology and Imaging Physics, The University of Texas M.D. Anderson Cancer Center, Houston, USA; Department of Neurology, Korea University Guro Hospital, Seoul, Republic of Korea; Department of Neurology, Seoul National University Bundang Hospital, Seongnam, Republic of Korea; Department of Neurology, Samsung Medical Center, Sungkyunkwan University, Seoul, Republic of Korea; Department of Neurology, Asan Medical Center, Seoul, Republic of Korea; Department of Neurology, Chonnam National University Hospital, Gwangju, Republic of Korea; Department of Neurology, Dong-A University Hospital, Busan, Republic of Korea; Department of Radiology, Seoul National University Bundang Hospital, Seongnam, Republic of Korea; Department of Radiology, Korea University Guro Hospital, Korea University College of Medicine, Seoul, South Korea; National Priority Research Center for Stroke, Goyang, Republic of Korea

**Author notes:** Correspondence to: Dong-Eog Kim, MD, PhD., Professor of Neurology, Dongguk University College of Medicine & Chair of National Priority Research Center for Stroke. Address: Department of Neurology, Dongguk University Ilsan Hospital, 27, Dongguk-ro, Ilsandong-gu, Goyang, South Korea.

## Abstract

**BACKGROUND:** Accurate classification of ischemic stroke subtype is important for effective secondary prevention of stroke. We used diffusion-weighted imaging (DWI) and atrial fibrillation (AF) data to train a deep learning algorithm to classify stroke subtype.

**METHODS:** Model training, validation, and internal testing were done in 2,988 patients with acute ischemic stroke from three stroke centers by using U-net for infarct segmentation and EfficientNetV2 for stroke subtype classification. Experienced vascular neurologists (n=5) determined stroke subtypes for external test datasets, while establishing a consensus for clinical trial datasets using the TOAST classification. Infarcts on DW images were automatically segmented using an artificial intelligence solution that we recently developed, and their masks were fed as inputs to a deep learning algorithm (DWI-only algorithm). Subsequently, another model was trained, with the presence or absence of AF included in the training as a categorical variable (DWI+AF algorithm). These models were tested: a) internally against the opinion of the labeling experts, b) against fresh external DWI data, and also c) against clinical trial DWI data acquired at a later date.

**RESULTS:** In the training-and-validation datasets, the mean age was 68.0±12.5 (61.1% male). In internal testing, compared with the experts, the DWI-only algorithm and the DWI+AF algorithm respectively achieved moderate (65.3%) and near-strong (79.1%) agreement. In external testing, both algorithms again showed good agreements (59.3-60.7% and 73.7-74.0%, respectively). In the clinical trial dataset, compared with the expert consensus, percentage agreements and Cohen’s kappa were respectively 58.1% and 0.34 for the DWI-only algorithm vs. 72.9% and 0.57 for the DWI+AF algorithm. The corresponding values between experts were comparable (76.0% and 0.61) to the DWI+AF algorithm.

**CONCLUSIONS:** Our deep learning algorithm trained on a large dataset of DWI (both with or without AF information) was able to classify ischemic stroke subtypes as accurately as a consensus of stroke experts.

## Introduction

Studies have shown that the volume^1^ and pattern^2^ of ischemic lesions on diffusion-weighted MRI (DWI) are associated with stroke subtype and predictive of post-stroke functional outcomes and future cerebrovascular events. Approximately a quarter of patients with ischemic stroke experience recurrence.^3,4^ In a previous study of 7,101 patients with acute ischemic stroke, we observed that large artery atherosclerosis (LAA) and cardio-embolic (CE) strokes were associated with a ∼5-times higher risk of recurrence at 1-year, compared with small vessel occlusion (SVO) stroke.^5^ The etiology of stroke is critical to the correct implementation of future preventative strategies.

The Trial of Org10172 in Acute Stroke (TOAST) classification has been the most frequently method employed for etiologic stroke subtyping in clinical practice and research.^6^ The original TOAST classification required clinical features and data from tests including brain imaging (CT/MRI), cardiac evaluation (electrocardiography, echocardiography, and etc.), duplex imaging of extracranial arteries, arteriography, and laboratory assessments for a pro-thrombotic state.^6^ Additional tests, such as Holter monitoring, implantable loop recorder, and high-resolution vessel wall MRI, have enabled more precise stroke subtyping.^7^ However, these tests increase the cost and the length of hospital stay. Moreover, many countries lack enough access to these advanced techniques. A diagnosis support system using initial or simple exams, such as DWI and electrocardiography (ECG), to detect acute infarcts and atrial fibrillation (AF) could reduce costs^8,9^ and assist clinicians who do not have access to other resources to determine stroke etiology.

To date, a few previous studies have developed automated systems for classifying stroke subtypes using deep learning algorithms and DWI.^10,11^ However, no study has externally validated these algorithms, which is critically important given the low inter-rater reliability in the classification of stroke subtypes.^12^ In the present multi-center study, we enrolled about 6,500 patients with acute ischemic stroke. Using 2,489 patients’ DWI data with and without information on the presence of AF, we developed a deep learning algorithm to classify stroke subtypes. We then externally validated the deep learning algorithm on a new set of 3,384 patients, using three temporally and regionally different datasets. In addition, we compared stroke subtype classifications by the deep learning algorithm vs. neurovascular experts. Finally, we outlined practical applications of the deep learning-based stroke subtype classification for cardioembolism risk stratification based solely on initial DWI assessments, for use when AF information is not available or becomes available after continuous ECG monitoring (for days ∼ years). ^13,14^

## Methods

### Participants

#### Datasets for training, validation, and internal testing

From May 2011 to March 2014, we consecutively enrolled 4,514 patients with acute ischemic stroke, who were admitted to three university hospitals (Dongguk University Hospital, Seoul National University Bundang Hospital, and Dong-A University Hospital) within 7 days of symptom onset. A total of 1,516 patients were excluded for the following reasons: (1) unavailable or poor-quality DWI (n = 342), (2) other causes of strokes such as arterial dissection, moyamoya disease, cancer-related stroke, etc. (n = 241), and (3) undetermined cause of stroke (n = 933). The remaining 2,998 patients’ data were used for training, validation, and internal test, using random sub-setting in ratio of 7:2:1 (Figure S1). The institutional review board of Dongguk University Hospital approved the study protocol (IRB No. 2017-09-017), and patients or their legal proxies provided a written informed consent.

#### Datasets for external testing

A total of 3,384 fresh stroke imaging datasets were used for external testing, comprised of the following components:

**External test dataset 1** From May 2011 to March 2014, 2,787 patients with acute ischemic stroke who were admitted within 7 days of symptom onset were consecutively enrolled from Chonnam National University Hospital. After excluding 868 patients, 1,919 were finally included.

**External test dataset 2** From October 2021 to August 2022, 1,315 patients with acute ischemic stroke who were admitted within 7 days of symptom onset were enrolled from the Chonnam National University Hospital. After excluding 491 patients, 824 were finally included.

**External test dataset 3** From March 2021 to April 2022, 931 patients with acute ischemic stroke who were admitted within 7 days of symptom onset were enrolled from Korea University Guro Hospital. After excluding 290 patients, 641 were finally included.

#### Clinical trial datasets

A pivotal clinical trial was conducted to assess the efficacy of deep learning algorithms in comparison to a standard reference established through expert consensus, and to measure the level of agreement between the deep learning algorithm and the consensus as well as among the experts themselves. From March 2016 to May 2017, 1,701 patients who met the following inclusion criteria were enrolled from the two stroke centers (Dongguk University Hospital and Seoul National University Bundang Hospital): 1) age between 20 and 95 years, 2) patients with acute ischemic stroke who visited the hospitals within 7 days after symptom onset, and 3) patients who underwent DWI. According to the prespecified exclusion criteria, we excluded 612 patients according to the following reasons: (1) inadequate or poor-quality DWI (n = 148), other causes of strokes (n = 114), and undetermined causes of strokes (n = 350), leaving 900 patients’ data for clinical testing.

### Clinical data collection

Using a standardized protocol,^15^ we prospectively collected demographic data, prior medication history, and the presence of vascular risk factors including hypertension, diabetes mellitus, hyperlipidemia, coronary artery disease, AF, and smoking history.

### Imaging acquisition and infarct segmentation

For the training data, brain MRIs were performed on 1.5 Tesla (n = 2,471) or 3.0 Tesla (n = 527) MRI systems. The DWI protocol was as follows: b-values of 0 and 1000 s/mm2, TE (echo time) 50-99 ms, TR (repetition time) 2400-9000 ms, voxel size 1×1×3∼5 mm^3^, interslice gap of 0–2 mm, and slice thickness of 3–7mm. Using a validated 3D U-net algorithm, we automatically segmented infarct lesions on DW images.^16,17^

#### Ischemic stroke subtype classification

For the datasets for training and validation, internal testing, and external test datasets 1-3, stroke subtypes were determined by experienced vascular neurologists at each hospital, using a validated MRI-based classification system built on the TOAST criteria (details provided in supplementary materials).^7^ Briefly, the modified TOAST classification is composed of the following five steps: 1) consideration of other determined etiologies of stroke; 2) screening for SVO on DWI; 3) consideration of relevant artery stenosis or occlusion; 4) consideration of recanalization status after thrombolytic therapy; and 5) consideration of follow-up recanalization status without thrombolytic therapy. For the clinical trial dataset, stroke subtypes were determined through consensus among three experienced vascular neurologists (J-.W.Chung, C.K.Kim, and D-.E.Kim).

#### Development of a deep learning algorithm for ischemic stroke subtype classification

Brain DWIs were preprocessed by (1) skull stripping using the Gaussian blur and Otsu’s threshold,^18^ (2) applying N4 bias field correction using the SimpleITK library, and (3) performing image signal normalization. After the preprocessing, infarct areas on DWI were automatically segmented using the validated 3D U-net algorithm.^16,17^ The segmented infarct masks from raw DW images were stacked and condensed into three 2D X, Y, Z-axis images to ensure consistent data input regardless of the number of slices (Figure S2). These condensed 2D X, Y, Z-axis images were resized to 256 x 256 pixels using bilinear interpolation. Thus, the training data for the algorithm was comprised of: three 2D images representing X, Y, Z-axis projections of segmented infarct area, and a label (LAA, SVO, and CE).

For the training, we utilized the EfficientNet v2,^19^ a new family of convolutional networks that have faster training speed and better parameter efficiency, while adding a global_average_pooling2d layer to minimize overfitting by reducing the total number of parameters. In addition, we incorporated a sequence of one inner dense layer with dropout layers. In total, a 30% dropout rate was randomly chosen to avoid overfitting. Finally, one output dense layer contained 3 output units for multi-class (LAA, SVO, and CE) classification, which were designated as the DWI-only based subtype classification. The details of the layers, their order in the proposed model, and the output shape of each layer are presented in Figure S2. The total number of parameters was 52,862,199.

To develop a deep learning algorithm that takes account for AF, we concatenated a binary value (0 vs. 1: the absence vs. presence of AF) to previous outputs, and then applied a fully connected layer. The output was then designated as DWI+AF based subtype classification.

For all the procedures, including preprocessing and model development, we used Python 3.7.9 and 3.8.13, PyTorch 1.12.0, Torchvision 0.13.0, pandas 1.2.4, NumPy 1.19.5/1.22.3, SciPy 1.4.1/1.6.3, scikit-image 0.15.0/0.18.1, SimpleITK 2.1.1, and Pydicom 2.1.2. Each model was trained for ∼9 hours using a hardware system comprising Intel Xeon Silver 4314 @2.40GHz, 640GB RAM, and NVIDIA Quadro RTX A6000 with 48GB GDDR6.

### Expert consensus for the classification of stroke subtype in the clinical trial dataset

For the clinical trial dataset, we first assessed the inter-observer agreement of stroke subtype classification between two experts (J-W Chung and J-S Lim, board-certified neurologists with more than 5-year experience in both stroke practice and research), who had served as stroke neurologists at least five years and independently reviewed the brain MRI and patients’ data. Information provided to the reviewers included age, sex, the presence of AF, DW images, and MR or CT angiography. Based on the aforementioned ischemic stroke subtype-classification system,^7^ they independently determined etiologies (i.e., LAA, SVO, or CE). In cases of disagreement between the two reviewers, a third reviewer (D-E Kim) served as the tie-breaker. When the final consensus on stroke subtype was undetermined or other determined stroke, the case was excluded from the analysis. The experts’ consensus classifications were compared with the deep learning algorithm’s classifications.

### Statistical analysis

The baseline characteristics between datasets were compared using the ANOVA or Kruskal-Wallis test for continuous variables and Chi-square test for categorical variables, as appropriate. To compare the subtype classifications made by experts and those made by deep learning algorithms, we used percentage agreement and Cohen’s kappa. In addition, we calculated the sensitivity, specificity, positive predictive value, and negative predictive value for each subtype (LAA, SVO, and CE). To examine the clinical implications of artificial intelligence (AI) prediction of cardioembolism using DWI, participants in each dataset were stratified into ten groups based on the probability of having cardioembolism estimated by deep learning algorithm. The trend of the observed frequency of cardioembolic stroke, as determined by experts, was examined using a Wilcoxon-type test for trend.^20^ All the statistical analyses described above were performed using STATA 16.0 (STATA Corp., Texas, USA), and a p-value < 0.05 was considered statistically significant.

## Results

### Baseline characteristics

In the training and validation datasets, the mean age was 68.0±12.5 and 61.1% were men (Table 1). Mean ages were similar in all datasets. Other demographic characteristics, such as sex, admission National Institute of Health Stroke Scale (NIHSS) scores, and risk factors for stroke, varied significantly among the datasets. The distribution of stroke subtypes also differed among the datasets, indicating their heterogeneity.

**Table 1.** Baseline characteristics.

|  | Training and validation (n = 2,687) | Internal test (n = 311) | External test dataset 1 (n = 1,919) | External test dataset 2 (n = 824) | External test dataset 3 (n = 641) | Clinical trial dataset (n = 900) | p value |
| --- | --- | --- | --- | --- | --- | --- | --- |
| Age | 68.0 ± 12.5 | 68.2 ± 12.9 | 68.7 ± 12.0 | 70.2 ± 12.4 | 67.5 ± 12.0 | 68.6 ± 12.4 | 0.23 |
| Sex, men | 1,642 (61.1%) | 174 (56.0%) | 1,090 (56.8%) | 504 (61.2%) | 426 (66.5%) | 551 (61.3%) | < 0.001 |
| Admission NIHSS score<sup>a</sup> | 4 (2 – 9) | 4 (2 – 9) | 4 (2 – 10) | 2 (1 – 5) | 3 (1 – 5) | 4 (2 – 7) | < 0.001<sup>b</sup> |
| Prestroke mRS score 0~2<sup>a</sup> | 2,346 (87.3%) | 274 (88.1%) | 1,667 (87.9%) | 791 (96.0%) | 126 (74.6%) | 832 (92.5%) | < 0.001 |
| Stroke subtype |  |  |  |  |  |  | < 0.001 |
| LAA | 1,224 (45.6%) | 142 (45.7%) | 1,044 (54.4%) | 434 (52.7%) | 300 (46.8%) | 574 (63.8%) |  |
| SVO | 667 (24.8%) | 75 (24.1%) | 221 (11.5%) | 154 (18.7%) | 222 (34.6%) | 155 (17.2%) |  |
| CE | 796 (29.6%) | 94 (30.2%) | 654 (34.1%) | 236 (28.6%) | 119 (18.6%) | 171 (19.0%) |  |
| Prior stroke history | 570 (21.2%) | 68 (21.9%) | 301 (15.7%) | 161 (19.5%) | 101 (15.8%) | 168 (18.7%) | < 0.001 |
| Coronary artery disease | 257 (9.6%) | 29 (9.3%) | 60 (3.1%) | 67 (8.1%) | 56 (8.7%) | 82 (9.1%) | < 0.001 |
| Hypertension | 1,884 (70.1%) | 222 (71.4%) | 1,203 (62.7%) | 484 (58.7%) | 426 (66.5%) | 660 (73.4%) | < 0.001 |
| Diabetes | 966 (36.0%) | 126 (40.5%) | 577 (30.1%) | 247 (30.0%) | 205 (32.0%) | 341 (37.9%) | < 0.001 |
| Hyperlipidemia | 1,246 (46.4%) | 148 (47.6%) | 299 (15.6%) | 154 (18.7%) | 68 (10.6%) | 385 (42.8%) | < 0.001 |
| Smoking | 1,122 (41.8%) | 121 (38.9%) | 717 (37.4%) | 219 (26.6%) | 215 (33.5%) | 394 (43.8%) | < 0.001 |
| Atrial fibrillation | 645 (24.0%) | 74 (24.0%) | 539 (28.1%) | 230 (27.9%) | 98 (15.3%) | 172 (19.1%) | < 0.001 |
| Recanalization therapy | 458 (17.1%) | 53 (17.0%) | 448 (23.4%) | 119 (14.4%) | 72 (11.2%) | 135 (15.0%) | < 0.001 |

<sup>a</sup>Data were missing in 472 and 1 patients of the external test dataset-3 and clinical trial dataset, respectively. <sup>b</sup>Kruskal-Wallis test was used. NIHSS=National Institute of Health Stroke Scale; mRS=modified Rankin Scale ; LAA=large artery atherosclerosis; SVO=small vessel occlusion; CE=cardioembolism.

### Deep learning prediction of stroke subtype using DWI data only vs. DWI plus AF data

In the internal test dataset (Figure 1), the percentage agreement between the DWI-only algorithm and stroke experts was 65.3% (95% CI: 60.0–70.6%). After incorporating the information regarding the presence of AF (DWI+AF algorithm), the percentage agreement was increased to 79.1% (95% CI: 74.6–83.6%).

**Figure 1.**
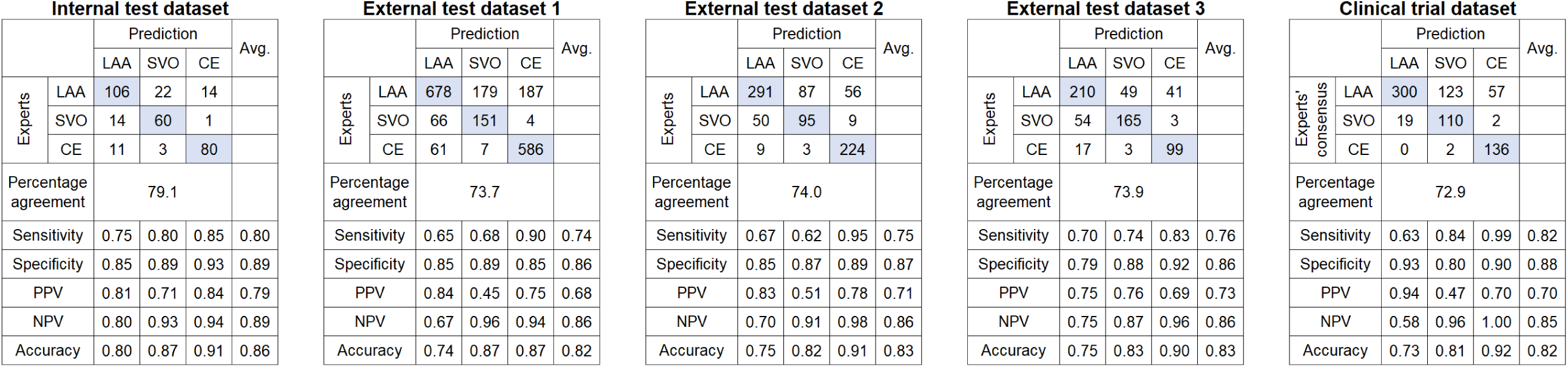
Confusion matrix for deep learning algorithm vs. expert classification of stroke subtype using diffusion-weighted images and atrial fibrillation information. For each stroke subtype, sensitivity, specificity, positive predictive value (PPV), negative predictive value (NPV), and accuracy were evaluated. The average value of each statistic was shown in the last column. LAA=large artery atherosclerosis; SVO=small vessel occlusion; CE=cardioembolism; Avg=average.

In the external test datasets (Figure 1), both algorithms again showed good agreements. The DWI-only algorithm achieved 58.1 ∼ 60.7% levels of agreements (Table 2, Figure S3). The DWI+AF algorithm again showed higher agreements, ranging from 73.7% to 74.0%, with Cohen’s Kappa ranging from 0.57 to 0.59. In addition, the accuracy of stroke subtype classification reached 0.83.

**Table 2.** Agreements of stroke subtype classification between deep learning algorithm and stroke neurologists (experts)

|  | Internal test<br>(n = 311) | External test dataset 1<br>(n = 1,919) | External test dataset 2<br>(n = 824) | External test dataset 3<br>(n = 641) | Clinical trial dataset (n<br>= 900) |
| --- | --- | --- | --- | --- | --- |
|  | Deep learning<br>algorithm vs. experts | Deep learning<br>algorithm vs. experts | Deep learning<br>algorithm vs. experts | Deep learning<br>algorithm vs. experts | Deep learning<br>algorithm vs. experts'<br>consensus |
| DWI-only algorithm |  |  |  |  |  |
| Percentage agreement (95%<br>CI) | 65.3% (60.0 – 70.6%) | 60.7% (58.5 – 62.9%) | 59.8% (56.5 – 63.2%) | 59.3% (55.5 – 63.1%) | 58.1% (54.9 – 61.3%) |
| Cohen's kappa (95% CI) | 0.47 (0.39 – 0.56) | 0.38 (0.34 – 0.41) | 0.37 (0.32 – 0.42) | 0.37 (0.31 – 0.43) | 0.34 (0.29 – 0.39) |
| DWI+AF algorithm |  |  |  |  |  |
| Percentage agreement (95%<br>CI) | 79.1% (74.6 – 83.6%) | 73.7% (71.8 – 75.7%) | 74.0% (71.0 – 77.0%) | 74.0% (70.5 – 77.4%) | 72.9% (69.1 – 76.7%) |
| Cohen's kappa (95% CI) | 0.68 (0.61 – 0.75) | 0.57 (0.54 – 0.60) | 0.59 (0.54 – 0.64) | 0.59 (0.54 – 0.64) | 0.57 (0.51 – 0.62) |
| Between experts |  |  |  |  |  |
| Percentage agreement (95%<br>CI) |  |  |  |  | 76.0% (74.0 – 79.0%) |
| Cohen's kappa (95% CI) |  |  |  |  | 0.61 (0.57 – 0.65) |

508 DWI=diffusion-weighted image; CI=confidence interval; AF=atrial fibrillation.

In the clinical trial dataset (Figure 1), the percentage agreements and Cohen’s kappa were respectively 58.1% (95% CI: 54.9–61.3%) and 0.34 (0.29–0.39) for the DWI-only algorithm, and the values were 72.9% (95% CI: 69.1–76.7%) and 0.57 (0.51–0.62) for the DWI+AF algorithm, respectively.

Alluvial plots for the five datasets (Figure 2) showed that additional information regarding the presence of AF on ECG changed the categorization of stroke subtype by the DWI-only algorithm from CE to LAA more often (22.1∼38.2%) than from LAA to CE (13.7∼16.2%) or from SVO to CE (4.2∼7.2%). There was no reclassification from CE to SVO.

**Figure 2.**
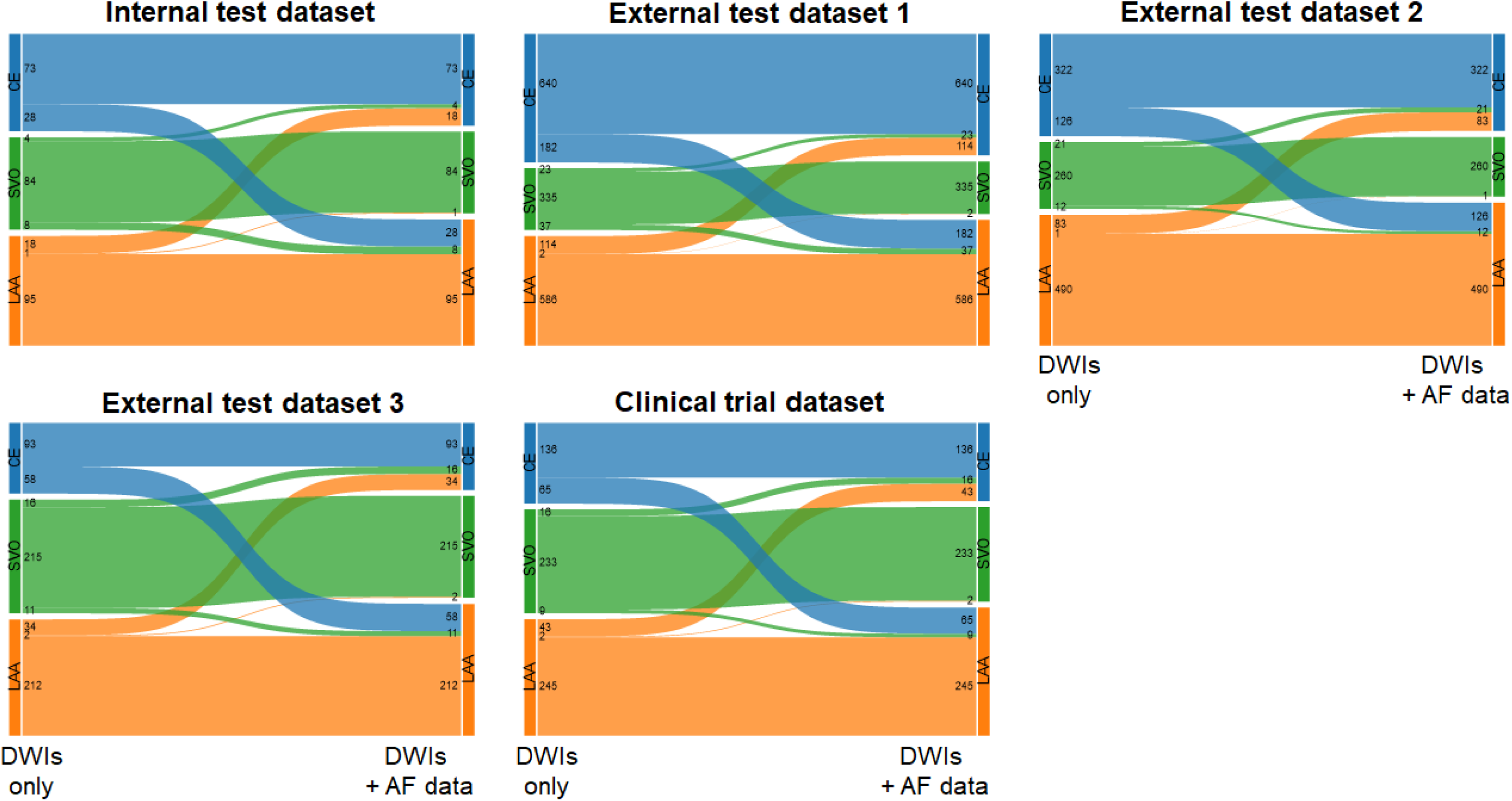
Alluvial plot depicting changes of stroke subtype classification after using atrial fibrillation (AF) data in addition to diffusion-weighted images (DWIs) Numbers indicates the number of patients in each stroke subtype. LAA=large artery atherosclerosis; SVO=small vessel occlusion; CE=cardioembolism.

### DWI-based prediction of cardioembolism

When we divided subjects into deciles of the expected CE probability (estimated by the DWI-only algorithm; Table S1), the observed frequency of the CE subtype determined by experts increased with a nearly linear fashion (Figure 3), showing good agreement. A similar trend was observed in all external test datasets. In the 8^th^, 9^th^, and 10^th^ decile groups, approximately 40-70% of subjects were shown to have CE strokes. Furthermore, in the clinical trial dataset, there was a strong correlation between the expected probability and observed frequency.

**Figure 3.**
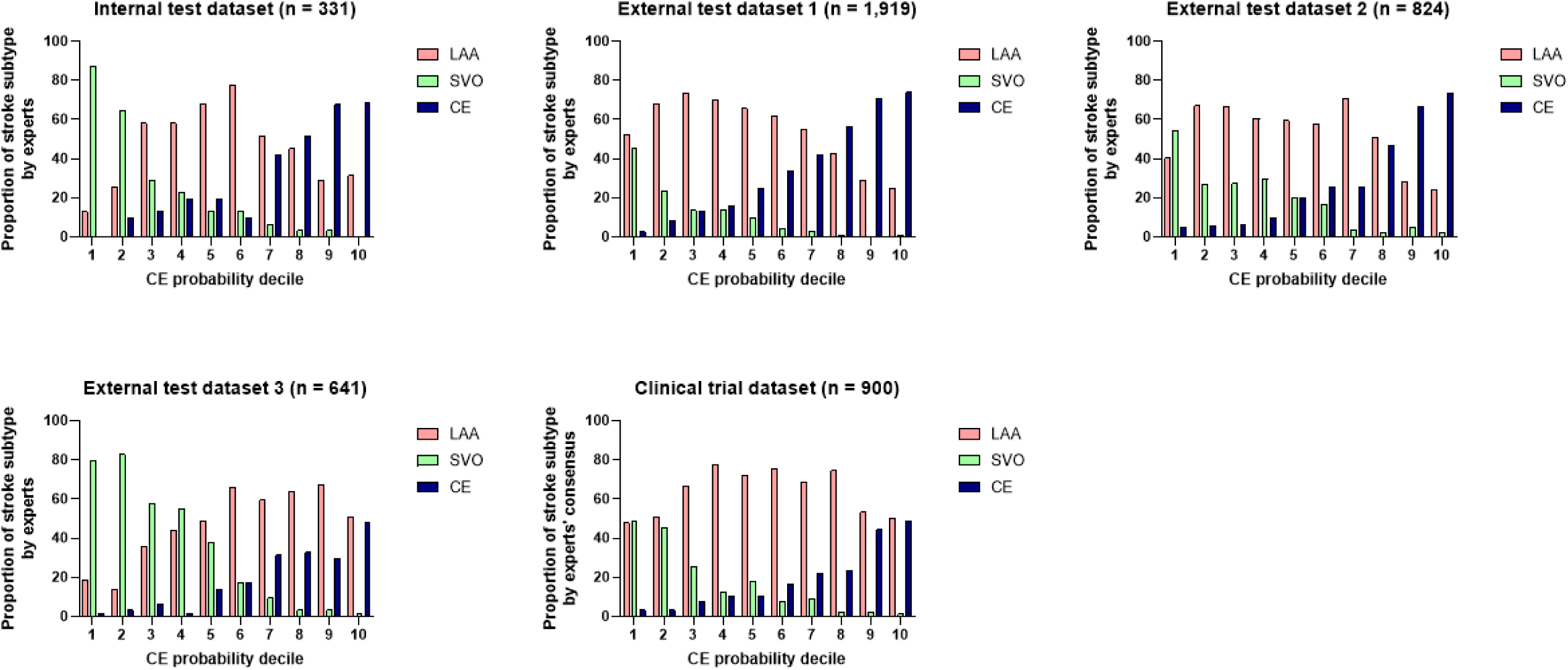
Proportions of stroke subtypes determined by experts in each decile of increasing cardioembolic (CE) probability that was estimated by the DWI-only based deep learning algorithm. Using diffusion-weighted images only, a deep learning algorithm estimated probabilities of CE stroke. Then, the probabilities of every case were categorized into deciles in each dataset. Bars indicate observed frequency of each stroke subtype determined by experts. Note that the proportion of CE stroke diagnosed rises proportionally with the estimated CE probability, suggesting that both human experts and the AI, are examining the same underlying entity. LAA=large artery atherosclerosis; SVO=small vessel occlusion.

## Discussion

In the present study, we developed a fully automated deep learning algorithm to classify ischemic stroke subtype using DWI and AF data from 2,998 ischemic stroke patients from three stroke centers. The deep learning algorithm was externally validated with three external datasets. The algorithm demonstrated good agreement with stroke experts, achieving Cohen’s kappa coefficients of 0.57∼0.59 for three external datasets, which were lower than the value (0.68) for the internal dataset. Furthermore, the clinical trial also demonstrated that the AI classification of stroke subtypes was comparable to the expert consensus.

To date, few studies have developed deep learning algorithms to classify stroke subtypes. According to a study that exclusively utilized electronic medical records, deep learning algorithms demonstrated moderate agreement (kappa = 0.57) when compared with expert decisions.^21^ Another study reported that a deep learning algorithm to classify stroke subtypes using DWI showed an average accuracy of 81.9%.^22^ However, these investigations did not conduct an external validation. As described, the present study validated our deep learning algorithm in three different external datasets and in a clinical trial involving two hospitals. This represents the largest dataset, and best external validation, currently available in the literature, to our knowledge. In all datasets, the deep learning algorithm achieved a similarly high mean accuracy (between 0.82 and 0.83), supporting its robustness. It is notable that there was a comparable level of agreement between the consensus of experts and deep learning predictions as there was between the experts themselves.

Studies have demonstrated that stroke subtypes are closely associated with the pattern and extent of ischemic lesions.^2,23,24^ Cardioembolic strokes were associated with corticosubcortical single lesions, multiple lesions in anterior and posterior circulations, and multiple lesions in multiple cerebral circulations (P = 0.008).^2^ LAA stroke lesions were located more frequently in the same vascular territory than CE strokes.^23, 24^ SVO stroke could be distinguished from other stroke subtypes based on distinctive morphological properties.^23^ Thus, our deep learning algorithm trained on extensive DWI data may infer morphological and geometrical patterns associated with stroke etiologies.

Guidelines for secondary prevention of stroke underscore a tailored therapeutic approach based on stroke subtypes,^25,26^ recommending strict blood pressure management for SVO strokes,^27^ intensive antiplatelet and lipid-lowering therapy for LAA strokes,^28–31^ and anticoagulant therapy for CE strokes.^32^ However, a quarter of strokes are classified as embolic stroke with undetermined source (ESUS).^33^ Repeated failures of randomized clinical trials (RCTs) to compare the effectiveness of antiplatelets and direct oral anticoagulants in preventing stroke in patients with ESUS^34–36^ have highlighted the need for new biomarkers or tools to identify people at high risk of CE stroke. A few machine learning algorithms using clinical and echocardiography data have demonstrated promising results in identifying individuals with an increased risk of AF within ESUS subjects.^33,37^ However, these algorithms relied on extensive data input such as patients’ demographics, vascular risk factors, comorbidities, vital signs, laboratory results, and echocardiographic findings. The comprehensive data requirement poses a challenge in real-world scenarios, where data acquisition varies and resources are often limited. Our deep learning algorithm identified CE strokes based solely on DWI, suggesting its potential clinical utility in predicting an occult cardioembolic source in ESUS without additional clinical and laboratory data.

In the CRYSTAL-AF (Cryptogenic Stroke [CS] and Underlying AF) trial, stroke was classified as cryptogenic when the cause remained uncertain after extensive diagnostic evaluation, including 12-lead ECG, 24 hours or more of ECG monitoring, transesophageal echocardiography, angiographic or ultrasonographic evaluation of intracranial and extracranial vessels, and screening for thrombophilic states (in patients <55 years of age). ^14^ In this study, ECG monitoring with an insertable cardiac monitor detected AF in 12.4% of patients by one year.^14^ We hypothesize that AI algorithms can increase the yield of testing, by helping to select patients who are more likely to test positive for AF during long-term ECG monitoring. To test the hypothesis, further research should investigate prospectively whether an occult cardioembolic source is more often found during post-ESUS or post-CS follow-up in patients with higher CE probabilities predicted by our DWI-only algorithm.

Including AF information changed the DWI-only algorithm-based original categorization of stroke subtype in about 20% of cases, which highlights the importance of detecting AF. In the NAVIGATE ESUS (New Approach Rivaroxaban Inhibition of Factor Xa in a Global Trial Versus ASA to Prevent Embolism in Embolic Stroke of Undetermined Source) trial, rivaroxaban failed to show superiority over aspirin in preventing recurrent ischemic stroke (4.7% per year in both groups).^35^ It was suggested that the eligibility assessment may not have effectively identified strokes due to embolism and that AF was not a major cause of recurrent stroke.^35,38^ Indeed, AF was identified in only 3% of the patients at a median follow-up of 5 months, although systematic screening for arrhythmia was not performed during the trial.^35^ However, the role of AF in patients with ESUS, whether it is the underlying cause of the index stroke or not, and its effect on stroke recurrence remain unclear,^39^ requiring further investigations. In the NAVIGATE ESUS trial, about two-thirds of carotid plaques were present in the carotid artery ipsilateral to the index stroke, showing a strong trend of a higher risk of recurrent ischemic stroke.^35^ Thus, future ESUS trials for direct oral anticoagulants may have to exclude strokes due to carotid atherosclerosis.^40^ Our deep learning algorithms, which effectively classify stroke subtypes using DW images with or without AF data, would facilitate these research, such as by improving eligibility assessments.

Our study has limitations. First, stroke experts typically determine ischemic stroke etiology by using clinical, angiographic, and laboratory data in a comprehensive manner. The validity of relying solely on DWI and AF information could be questioned. An earlier study demonstrated that TOAST diagnoses without DWI matched final diagnoses in 48%, improving to 83% after DWI alone and to 94% after DWI plus MRA,^41^ indicating that DWI features has a major impact on classification accuracy enhancement. Second, although we validated the algorithm using multiple external datasets of Korean stroke populations, further investigation is required for multi-ethnic populations.

In conclusion, our deep learning algorithm trained on a large dataset of DWI and AF information was able to classify ischemic stroke subtypes as accurate as stroke experts. The AI algorithm, which performed well with the minimal data input in three different external test datasets and a multi-center clinical trial dataset, could be useful for stroke management by less experienced physicians or general practitioners.

## Data Availability

Data generated or analyzed during the study are available from the corresponding author by request.

## Author Contribution

W-.S.Ryu, and D-.E. Kim had full access to all study data and take responsibility for data integrity and the accuracy of the data analysis. W-.S.Ryu, D.S. and D-.E. Kim contributed study’s concept and design and drafted the manuscript. K.-J.Lee, C.K.Kim, B.J.Kim, J-.W.Chung, J-.S.Lim, J-.T.Kim, D-.H.Kim, and J-.K.Cha contributed to the acquisition, analyzing, and interpretation of data. W-.S.Ryu contributed statistical analysis and data visualization. All authors contributed to the critical revisions of the manuscript for important intellectual content.

## Acknowledgements

The authors appreciate the contributions of all members of the Comprehensive Registry Collaboration for Stroke in Korea to this study. This study was supported by the Multiministry Grant for Medical Device Development (KMDF_PR_20200901_0098), the National Priority Research Center Program Grant (NRF-2021R1A6A1A03038865), and the Basic Science Research Program Grant (NRF-2020R1A2C3008295) of National Research Foundation, funded by the Korean government.

## Non-standard Abbreviations and Acronyms

DWI: diffusion-weighted
MRI LAA: large artery atherosclerosis
CE: cardioembolism
SVO: small vessel occlusion
TOAST: The Trial of Org10172 in Acute Stroke
ECG: electrocardiography
AF: atrial fibrillation
AI: artificial intelligence
NIHSS: National Institute of Health Stroke Scale
ESUS: Embolic Stroke with Undetermined Source
RCT: randomized clinical trial

